# Evaluation of saliva molecular point of care for detection of SARS-CoV-2 in ambulatory care

**DOI:** 10.1101/2021.06.12.21258811

**Authors:** Jérôme LeGoff, Solen Kernéis, Caroline Elie, Séverine Mercier Delarue, Nabil Gastli, Laure Choupeaux, Jacques Fourgeaud, Marie-Laure Alby, Pierre Quentin, Juliette Pavie, Patricia Brazille, Marie Laure Néré, Marine Minier, Audrey Gabassi, Chrystel Leroy, Béatrice Parfait, Jean-Marc Tréluyer, Constance Delaugerre

**Author notes:** **Corresponding author** Jerome Le Goff, Virologie, Hôpital Saint-Louis, 1 avenue Claude Vellefaux, 75010 Paris, France. Contributed equally (Solen Kernéis, Caroline Elie). Contributed equally (Jean-Marc Tréluyer, Constance Delaugerre).

## Abstract

**Background:** Rapid identification of SARS-Cov-2 infected individuals is a cornerstone in strategies for the control of virus spread. The sensitivity of SARS-CoV-2 RNA detection by RT-PCR is similar in saliva and nasopharyngeal swab. Rapid molecular point-of-care tests in saliva could facilitate, broaden and speed up the diagnosis.

**Objectives and methods:** We conducted a prospective study in two community COVID-19 screening centers to evaluate the performances of a CE-marked RT-LAMP assay (EasyCoV™) specifically designed for the detection of SARS-CoV2 RNA from fresh saliva samples, compared to nasopharyngeal RT-PCR (reference test), to saliva RT-PCR and to nasopharyngeal antigen testing.

**Results:** Overall, 117 of the 1718 participants (7%) were tested positive with nasopharyngeal RT-PCR. Compared to nasopharyngeal RT-PCR, the sensitivity and specificity of the RT-LAMP assay in saliva were 34% (95%CI: 26-44) and 97% (95%CI: 96-98) respectively. The performance was similar in symptomatic and asymptomatic participants. The Ct values of nasopharyngeal RT-PCR were significantly lower in the 40 true positive subjects with saliva RT-LAMP (Ct 25.9) than in the 48 false negative subjects with saliva RT-LAMP (Ct 28.4) (p=0.028). Considering six alternate criteria for reference test, including saliva RT-PCR and nasopharyngeal antigen, the sensitivity of saliva RT-LAMP ranged between 27 and 44%.

**Conclusion:** In the ambulatory setting, the detection of SARS-CoV-2 from crude saliva samples with the RT-LAMP assay had a lower sensitivity than nasopharyngeal RT-PCR, saliva RT-PCR and nasopharyngeal antigen testing.

**Registration number:** NCT04578509

**Funding Sources:** French Ministry of Health and the Assistance Publique-Hôpitaux de Paris Foundation.

## Introduction

Coronavirus disease 2019 (COVID-19) pandemic has caused significant impact on the healthcare system and socioeconomic activity. Early diagnosis is critical for prompt actions on patient management, infection control, and public health control measures (1). Since transmission can occur from asymptomatic or pre-symptomatic patients, mass testing, together with rigorous contact tracing and isolation, has been recommended to control the pandemic (2–4). This strategy implies rapid and reliable testing methods. Although molecular detection of SARS-CoV-2 RNA in nasopharyngeal swabs is considered as the “gold standard” for identifying infected individuals(1,5), nasopharyngeal sampling requires specific sampling equipment and trained personnel and may be difficult in some patients. Mass RT-PCR testing is carried out in specialized laboratories and needs several hours before results release. Altogether, these constrains restrain access to massive testing, increase time-to-result and consequently delay isolation of contagious individuals (6).

Rapid antigen point-of-care (Ag) testing allows to overcome the drawback of RT-PCR time-to-result but still requires nasopharyngeal sampling. Sensitivity of Ag tests was estimated at 50-90% and specificity at 90-100% as compared to nasopharyngeal RT-PCR (7,8). Recently self-anterior nasal sampling has been tested to reduce patient discomfort and avoid requirements for nasopharyngeal swabbing (9,10).

Self-collected saliva is non-invasive and easy to collect and thus more suitable for mass-screening than nasopharyngeal sampling (11–14). Recent meta-analyses assessed performances of saliva RT-PCR tests for the diagnosis of COVID-19 (15–18) and we recently confirmed in a large prospective study the excellent sensitivity of saliva RT-PCR, as compared to nasopharyngeal RT-PCR, for the detection of SARS-CoV-2 in community screening centers (19).

The combination of saliva sampling with rapid point-of-care testing could facilitate screening and isolation of infected individuals. Rapid single use RT-PCR assays for SARS-CoV2 RNA detection are available but were validated mainly on nasopharyngeal samples (20,21) and rarely in saliva (22), require sophisticated equipment and remain expensive. Nucleic acid detections based on isothermal amplification, such as loop-mediated isothermal amplification (LAMP), are interesting approaches as they simplify the analytical process, reduce the cost and enable to speed up the diagnosis. The sensitivity of RT-LAMP directly from NPS samples varies from 65% to 87 % compared to RT-PCR. Few studies tested RT-LAMP on self-collected saliva without RNA extraction. Sensitivities ranged from 45 to 85%, results being better after RNA purification than from crude samples (23–26). No studies estimated the performances of RT-LAMP on saliva samples as point-of-care systems directly in screening centers.

We conducted a prospective study in two community COVID-19 screening centers to evaluate the performances of a CE-marked RT-LAMP assay specifically designed for the detection of SARS-CoV2 RNA from fresh saliva samples compared to nasopharyngeal RT-PCR, saliva RT-PCR and nasopharyngeal Ag tests.

## Methods

### Study population and procedures

All adults and children, either symptomatic or asymptomatic, referred to the two participating COVISAN centers, Paris, France, were eligible as described previously (19). In accordance with EasyCoV® assay manufacturer instructions of (SkillCell-Alcen, Jarry, France), performances of saliva RT-LAMP were estimated on saliva tested in screening centers, immediately after collection (< 5 minutes) or stored immediately at 4°C and then tested within a maximal 90 minutes interval after collection. In addition, patients should have a valid nasopharyngeal SARS-CoV-2 RT-PCR test. Eligible persons received oral and written detailed information, adapted to their age. Data on sociodemographics, past medical history, presence of symptoms, consumption of alcohol, coffee, food, smoking and teeth brushing in the hours before testing were collected. The NPS was sent to the APHP high throughput platform for RT-PCR as part of routine care (reference method). Participants were asked to self-collect saliva sample after salivating 30 second in their mouth. Saliva were tested directly in the screening center (see below) and then centralized for RT-PCR testing and frozen at -80°C within 24 hours.

### Virology methods

#### Nasopharyngeal RT-PCR

NPS were centralized and processed according to the routine procedure ((19), appendix). Nucleic acid extraction was performed with MGIEasy Nucleic Acid Extraction Kit (MGI Tech Co, Shenzhen, China) on a MGISP-960 instrument (MGI Tech Co). SARS Cov-2 RNA amplification was done using TaqPath™ COVID 19 CE IVD RT PCR Kit (Thermo Fisher Scientific, Coutaboeuf, France). The technique provides results expressed as a cycle threshold (Ct) for each gene target (ORF1ab, N and S-genes). The cutoff value of RT-PCR cycle threshold (Ct) retained to distinguish high/significant and moderate/low SARS-coV2 loads with TaqPath™ COVID 19 CE IVD RT PCR Kit was 28 (27). Ct values equal or higher than 32 corresponded to low viral loads.

#### Saliva RT-PCR

Saliva samples were tested at the APHP high throughput platform with RT-PCR on MGI instrument as described previously ((19), appendix). A 300 µl aliquot of saliva was mixed with 300 µl of NucliSENS® lysis buffer (Biomerieux, Marcy l’Etoile, France). Nucleic acid extraction and SARS Cov-2 RNA amplification were performed with the same procedure used for nasopharyngeal RT-PCR.

#### Saliva RT-LAMP

The test EasyCov® (SkillCell-Alcen, Jarry, France) is a CE-marked extraction-free RT-LAMP test specifically developed for saliva samples as point of care (saliva POC-LAMP). Detection of SARS-CoV2 was carried out according manufacturer’s instructions (EasyCOV®, SkillCell) (appendix). The procedure includes a step of virus inactivation and lysis at 80 ° C for 10 minutes and a step of viral genome amplification at 65 ° C for 30 minutes. The two steps take place in the Easyvid® system. After amplification, a reagent sensitive to pH is added to reveal the amplification. The result is immediately read by visual observation. The color turns yellow for a sample positive for SARS-CoV2 RNA and remains orange for a sample negative for SARS-CoV2 RNA.

#### Nasopharyngeal rapid antigen test

Nasopharyngeal Ag testing was performed with Standard Q COVID-19 Ag test (SD Biosensor®, Chuncheongbuk-do, Republic of Korea). Standard Q COVID-19 Ag test is a chromatographic immunoassay for the detection of SARS-CoV-2 nucleocapsid (N) antigen. The result was read after 15 to 30 minutes according to instructions of the manufacturer.

#### Statistical Analysis

Sample size was calculated assuming that the sensitivity of the index tests was equal or superior to 60%. To allow sufficient precision (± 10%), 93 subjects with positive nasopharyngeal RT-PCR were needed in each of the two subgroups (symptomatic and asymptomatic participants). As preliminary results indicates that viral load were not different between symptomatic and asymptomatic patients, the scientific committee of the study, during a planned meeting on 16 December 2020, recommended to perform the analysis as soon as 93 subjects with positive nasopharyngeal RT-PCR were included, whether symptomatic or asymptomatic.

RT-PCR results were considered positive if at least one gene was detected. Analyses of tests results were carried out blind of the result of the others and of the participant’s clinical data. For RT-PCR technique, Ct values reported are those for the ORF1a gene, and if not amplified, of the N-gene for (and of S-gene if the N-gene was not amplified).

Quantitative data were expressed as median [interquartile range], and qualitative data as counts (percentages). Diagnostic accuracy of the index tests was evaluated by calculating sensitivity and specificity. Confidence intervals were calculated by the exact binomial method. Subgroups analyses were performed according to: i) the presence of symptoms on day of testing, ii) the Ct value of the nasopharyngeal RT-PCR, expressed as low (at least one of the 3 targets with Ct ≤ 28, i.e. high viral shedding), or high (all 3 targets with Ct > 28, i.e. low viral shedding), and iii) to the consumption of alcohol, coffee, food, and smoking or teeth brushing before sample collection.

Sensitivity analyses were performed considering 6 alternate criteria for positivity for the reference standard: i) ≥ 2 positive targets with nasopharyngeal RT-PCR, ii) ≥ 1 positive target with nasopharyngeal RT-PCR and at least one of the 3 targets with Ct < 32, iii) ≥ 1 positive target with saliva RT-PCR, iv) ≥ 1 positive target with either the nasopharyngeal or saliva RT-PCR, v) ≥ 1 positive target with either the nasopharyngeal or saliva RT-PCR and at least one of the 3 targets with Ct < 32, and vi) NPS antigen test.

Quantitative variables were compared with Wilcoxon’s test, with a significance level of 5%. The statistical analysis was performed using R software (http://cran.r-project.org/). Reporting of results followed the Standards for Reporting Diagnostic accuracy studies (STARD 2015) guideline (28).

### Institutional Review Board (IRB) approval

The IRB Ile-de France III approved the study protocol prior to data collection (approval number 3840-NI) and all subsequent amendments.

## Results

### Participants

Between November 4^th^ 2020 and February 15^th^ 2021, 1718 participants were enrolled with a nasopagryngeal sampling for RT-PCR and saliva sampling for RT-LAMP assay. Details of samples collected and tests performed for nasopharyngeal antigen assay and saliva RT-PCR are detailed in Figure 1. Median age of study participants was of 37 years [26-52] and 55% were females (Table 1). Indications for testing and clinical symptoms reported on day of inclusion are detailed in table 1. One to three symptoms were observed in 530/1712 (31%) participants.

**Table 1:** Characteristics of study participants. Results are presented as N (%) or median [interquartile range].

|  | Total<br>N=1718 |
| --- | --- |
| Age, years | 37 [26-52] |
| Females | 944 (55) |
| Contact with a confirmed case | 548 (32) |
| Time from last contact, days | 6 [1-7] |
| Presence of symptoms on day of testing | 691 (40) |
| Time from symptoms onset, days | 3 [2-4] |
| Cough | 329 (19) |
| Headaches | 268 (16) |
| Rhinorrhea | 264 (15) |
| Asthenia | 215 (13) |
| Muscle pain | 192 (11) |
| Fever | 163 (9) |
| Diarrhea | 107 (6) |
| Chills | 88 (5) |
| Anosmia | 46 (3) |
| Shortness of breath | 53 (3) |
| Chest pain | 50 (3) |
| Smoking in the last 24 hours | 331 (19) |
| Consumption of alcohol in the last 24 hours | 371 (22) |
| Consumption of coffee in the last hour | 347 (20) |
| Teeth brushing in the last 2 hours | 736 (43) |
| Mouth washing in the last 2 hours | 61 (4) |

**Figure 1.**
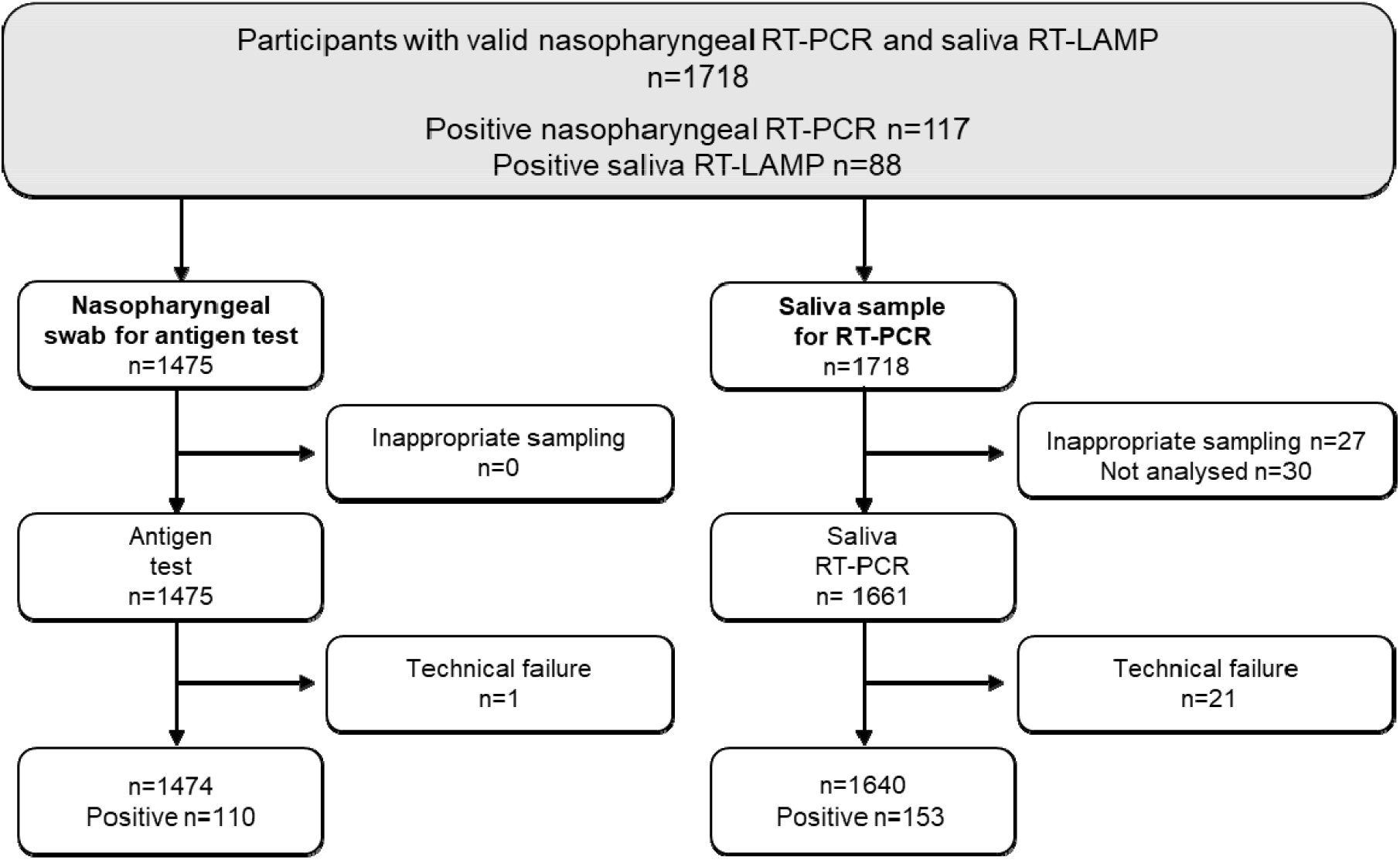
Study flowchart.

### SARS-CoV-2 positive results

Overall, 117/1718 (7%) tested positive on nasopharyngeal RT-PCR: 78/691 (11%) in symptomatic and 39/1027 (4%) in asymptomatic participants (Table 2). Detection rates were of 2%, 9% and 7% for saliva RT-LAMP, saliva RT-PCR and nasopharyngeal Ag test, respectively.

**Table 2:** Number of positive samples according to the technical procedure: nasopharyngeal RT-PCR, saliva RT-LAMP and RT-PCR, and nasopharyngeal antigen test.

|  | Positive/Total (%) | Presence of symptoms on day of testing |  |
| --- | --- | --- | --- |
|  |  | Symptoms | No symptoms |
| Nasopharyngeal RT-PCR | 117/1718 (7%) | 78/691 (11%) | 39/1027 (4%) |
| Saliva RT-LAMP | 88/1718 (5%) | 47/691 (7%) | 41/1027 (4%) |
| Saliva RT-PCR | 153/1640 (9%) | 93/662 (14%) | 60/978 (6%) |
| Nasopharyngeal antigen test | 110/1474 (7%) | 78/652 (12%) | 32/822 (4%) |

### Performance of detection of SARS-CoV-2 infection

Diagnostic accuracy of the two methods on saliva and the nasopharyngeal Ag test are presented in Table 3. Compared to RT-PCR on NPS, the sensitivity of saliva RT-LAMP was 34% (95% Confidence Interval (95%CI): 26-44). The sensitivity of saliva RT-PCR was 93% (95%CI: 86-97) and those of nasopharyngeal Ag test was 85% (95%CI: 77-91). The sensitivity and specificity of saliva RT-LAMP were similar in symptomatic and asymptomatic participants. Sensitivity analyses of saliva RT-LAMP according to six references (Table 4) showed similar results to the main analysis. Sensitivities of saliva RT-LAMP ranged between 27 to 44%. Its sensitivity was 37% (95%CI: 28-47%) compared to nasopharyngeal antigen test, and 30% (95%CI: 23-38) compared to saliva RT-PCR. Saliva RT-LAMP sensitivity was 40% (95%CI: 28-53) for Ct values below or equal to 28 and of 26% (95%CI: 15-40%) for Ct values above 28. As displayed on Figure 2, Ct values of nasopharyngeal RT-PCR were significantly lower in the 40 true positive subjects with saliva RT-LAMP (25.9 [19.4-30.2]) than in the 48 false negative subjects with saliva RT-LAMP (28.4 [24.4-32.6], p=0.028), with nasopharyngeal RT-PCR as reference test.

**Table 3:** Diagnostic accuracy of the saliva RT-LAMP and RT-PCR, and the nasopharyngeal antigen test as compared to the reference standard (nasopharyngeal RT-PCR, positivity defined as at least one target gene detected), according to the presence of symptoms in study participants.

|  | Total, n | Positive<br>samples, n | Sensitivity<br>(95% CI*) | Specificity<br>(95% CI) |
| --- | --- | --- | --- | --- |
| Saliva RT-LAMP | 1718 | 40 | 34% (26-44) | 97% (96-98) |
| Symptoms | 691 | 25 | 32% (22-44) | 96% (95-98) |
| No symptoms | 1027 | 15 | 38% (23-55) | 97% (96-98) |
| Saliva RT-PCR | 1640 | 153 | 93% (86-97) | 97% (96-97) |
| Symptoms | 662 | 93 | 93% (85-98) | 96% (94-97) |
| No symptoms | 978 | 60 | 92% (78-98) | 97% (96-98) |
| Nasopharyngeal antigen test | 1474 | 110 | 85% (77-91) | 99% (98-99) |
| Symptoms | 652 | 78 | 90% (81-96) | 98% (96-99) |
| No symptoms | 822 | 32 | 74% (57-88) | 99% (98-100) |

\*95% CI: 95% Confidence Interval

**Table 4:** Sensitivity analysis of diagnostic accuracy of the saliva RT-LAMP test, as compared to the several references.

| Reference standard | Total,<br>n | Positive<br>samples, n | Sensitivity<br>(95% CI*) | Specificity<br>(95% CI) |
| --- | --- | --- | --- | --- |
| NPS RT-PCR $\geq 2$ targets | 1718 | 88 | 35% (26-45) | 97% (96-98) |
| Symptoms | 691 | 47 | 34% (24-46) | 96% (95-98) |
| No symptoms | 1027 | 41 | 36% (21-54) | 97% (96-98) |
| NPS RT-PCR $\geq 1$ target and Ct value $<32$ | 1718 | 88 | 37% (27-47) | 97% (96-98) |
| Symptoms | 691 | 47 | 36% (24-49) | 96% (94-98) |
| No symptoms | 1027 | 41 | 38% (22-56) | 97% (96-98) |
| Saliva RT-PCR $\geq 1$ target | 1640 | 85 | 30% (23-38) | 97% (96-98) |
| Symptoms | 662 | 45 | 28% (19-38) | 96% (95-98) |
| No symptoms | 978 | 40 | 33% (22-47) | 98% (97-99) |
| NPS RT-PCR $\geq 1$ target or<br>Saliva RT-PCR $\geq 1$ target | 1648 | 87 | 28% (22-36) | 97% (96-98) |
| Symptoms | 667 | 47 | 27% (19-37) | 97% (95-98) |
| No symptoms | 981 | 40 | 30% (20-43) | 98% (97-99) |
| NPS RT-PCR $\geq 1$ target or<br>Saliva RT-PCR $\geq 1$ target and Ct value $<32$ | 1646 | 87 | 34% (26-42) | 97% (96-98) |
| Symptoms | 666 | 47 | 30% (21-41) | 97% (95-98) |
| No symptoms | 980 | 40 | 40% (26-55) | 98% (97-99) |
| NPS antigen | 1474 | 79 | 37% (28-47) | 97% (96-98) |
| Symptoms | 652 | 45 | 35% (24-46) | 97% (95-98) |
| No symptoms | 822 | 34 | 44% (26-62) | 97% (96-98) |

\*95% CI : 95% Confidence Interval.

**Figure 2.**
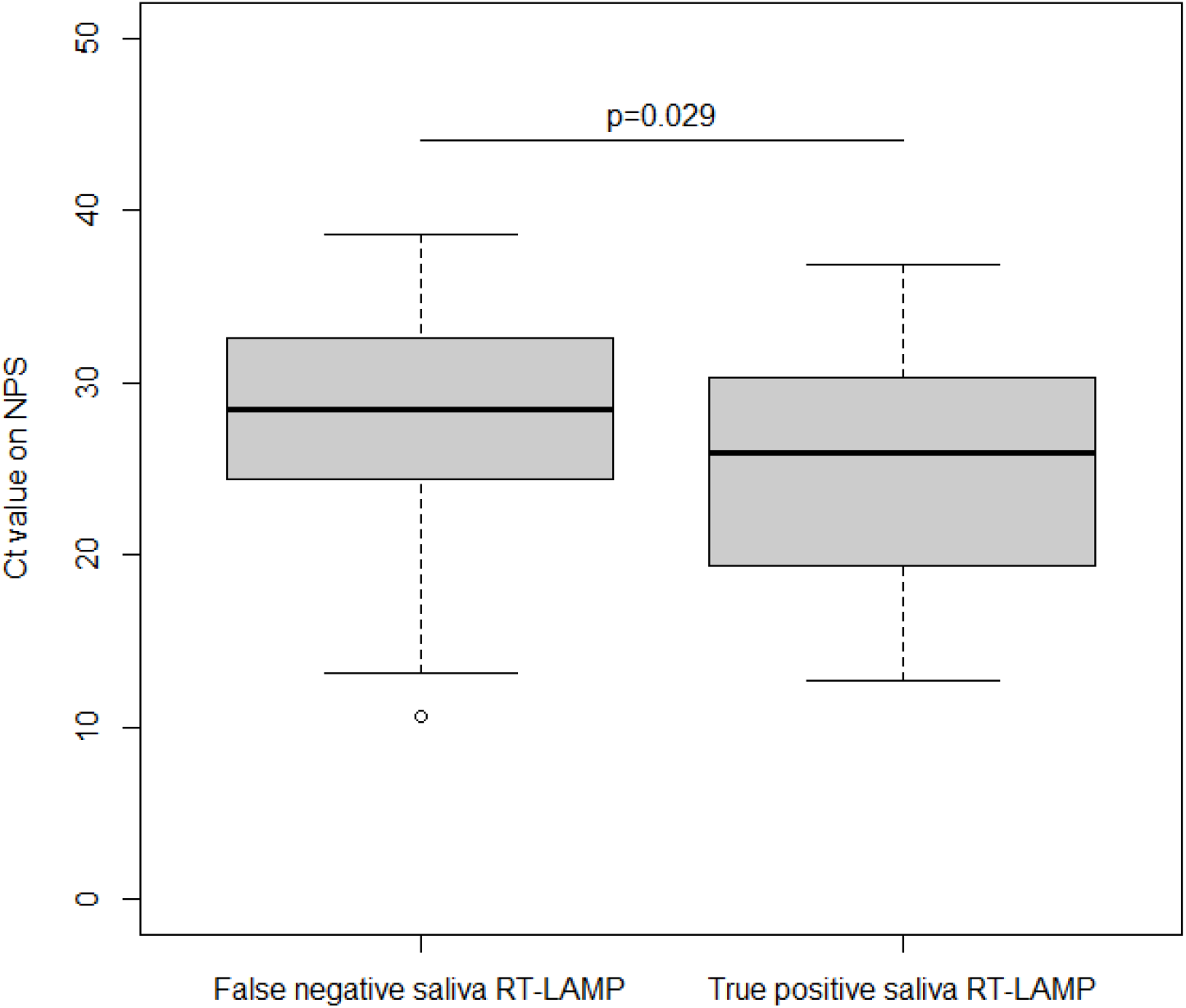
Nasopharyngeal SARS-CoV-2 RT-PCR Ct values according to saliva RT-LAMP results. Ct values of SARS-CoV-2 RT-PCR on nasopharyngeal samples (NPS) in individuals with saliva samples negative with the RT-LAMP assay (False negative) and those with saliva samples positive with the RT-LAMP assay (True positive) are presented in boxplots.

## Discussion

In this large prospective controlled study, the performance of a rapid RT-LAMP assay performed with crude saliva samples directly after saliva collection was analyzed. We used a CE marked assay specifically designed for saliva samples and for a point-of-care use. The test was authorized in France on November 2020 in symptomatic individuals for whom nasopharyngeal sampling was impossible or difficult. Our results showed, in a real-life rigorous evaluation, a low sensitivity of this method (34%) compared to nasopharyngeal RT-PCR. Its sensitivity remained low whatever the reference test considered (saliva RT-PCR, nasopharyngeal Ag test), ranging between 28 to 37%.

Our results differed strongly with the sensitivity of 86% (95CI 78%-94%) reported in Santos Schneider et al. study (29), and whatever the reference test used, nasopharyngeal RT-PCR or any other composite reference test including saliva RT-PCR and antigen test. In Santos Schneider et al. study, the authors evaluated the EasyCOV® assay in a central laboratory and tested each sample in triplicate. A sample was considered positive if at least two replicates out of three were positive. In our study, we tested all samples once directly in screening centers and according to manufacturer instructions, and to its expected use in routine conditions.

No difference in RT-LAMP sensitivity between symptomatic or asymptomatic participants was reported. Median time of testing was 3 days after symptom onset or 6 days after last contact of confirmed case. In 103 subjects already diagnosed for COVID-19, Nagura-Ikeda et al. reported, with another RT-LAMP assay, sensitivity results on saliva that differed according to the clinical state (23). The RT-LAMP assay was performed with nucleic acid extract of saliva instead of crude saliva. Overall sensitivity was of 71% compared to nasopharyngeal RT-PCR, with a higher sensitivity (85%) in patients tested within 9 days after symptom onset than after 9 days (44%). In asymptomatic individuals, the sensitivity was 60%.

According to other studies evaluating RT-LAMP tests, the critical step for sensitivity seemed to be the RNA extraction (13,23,25,30). A high level of concordance between RT-PCR on nasopharyngeal samples and RT-LAMP on saliva was observed when an automated extraction step (i.e. Qiasymphony RNA kit) was used. In a limited series of 34 positive samples (17 nasopharyngeal swabs and 17 saliva) tested by RT-PCR, Taki *et al*. reported a sensitivity of a RT-fluorescence LAMP assay performed with nucleic acid extracts of 97% and 100% in nasopharyngeal and saliva samples, respectively (25). Without RNA extraction on the same samples sensitivities decreased respectively to 71% and 47%, suggesting that RNA extraction process may be critical for the SARS-CoV-2 RNA detection by RT-LAMP especially for saliva samples. In our study, the RT-LAMP assay is an extraction-free test based on a 10 minutes heating at 80°C for virus inactivation and viral RNA release. This quick step suitable for a point-of-care test might be not optimal for RT-LAMP reaction with saliva samples and results may depend on miscellaneous factors according to the quality of saliva (volume, pH, viscosity, food by-products). The participants did not drink, eat or smoke within 30 minutes before saliva sampling. In addition, we did not find any significant effect of cigarettes or alcohol consumption within 2 or 24 hours. Another hypothesis is the impact of viral load. As we showed, Ct values of nasopharyngeal RT-PCR were lower in RT-LAMP true positive samples than in RT-LAMP false negative samples (26 vs 28), suggesting an impact of viral load on saliva RT-LAMP efficacy. However when considering only high or significant SARS-CoV-2 loads in nasopharyngeal samples, saliva RT-LAMP sensitivity remained low. Thus, the viral load per se does not explain the weak performance of the assay.

Finally, our study confirmed, as previously (19), the good performance of saliva RT-PCR and nasopharyngeal antigen testing as reliable alternative strategies to detect SARS-CoV-2 in both symptomatic and asymptomatic individuals in the ambulatory setting. Further work is needed to optimize an assay combining collected saliva and rapid point-of-care isothermal detection of SARS-CoV-2 RNA.

## Data Availability

Individual data will not be publicly available, because participants did not provide consent for it.

## Acknowledgments

The authors thank the following members of the scientific committee for insightful comments on the study protocol and results: Bruno Lina, Anne-Geneviève Marcelin, Catherine Paugam-Burtz, Astrid Vabret, Etienne Voirin-Mathieu, Michel Vidaud, Claire Poyart and staff involved in data collection: Claire Kappel, Guiseppina d’Anna, Elisabeth Velin, Abdulai Jalloh, Caroline Du Song, Philippine Treluyer, Mathilde Bayle, Lucie Daveau, Marine Hellegouarch, Suzie Zhu, Pénélope Travailleur, Matthieu Kapry, Adeline Huet, Aurélien Gibaud, Emeline Hermel, Céline Goy, Louise Lavancier.

### Appendix

#### SARS-CoV-2 RT-PCR on nasopharyngeal samples

NPS were inactivated at 56°C for 30 minutes. Nucleic acid extraction was performed on 180 µL of NPS with MGIEasy Nucleic Acid Extraction Kit (MGI Tech Co, Shenzhen, China) on a MGISP-960 instrument (MGI Tech Co). SARS-CoV-2 RNA amplification was done using TaqPath™ COVID 19 CE IVD RT PCR Kit (Thermo Fisher Scientific, Coutaboeuf, France). This test is a multiplex real-time RT-PCR test intended for the qualitative detection of nucleic acid from SARS-CoV-2. The kit contains three primer/probe sets specific to three different SARS-CoV-2 genomic regions (ORF1ab, N and S-genes) and primers/probes for bacteriophage MS2 used as internal control of amplification and extraction. A minimum of one negative control and one positive control was use for each run. Dilution of inactivated SARS-CoV-2 cell-culture supernatant (provided by Virology laboratory, Hospices Civils de Lyon) was added once a day in each production line as for external control. The technique provides results expressed as a cycle threshold (Ct) for each gene target.

#### SARS-CoV-2 RT-PCR on saliva samples

Saliva aliquots, stored frozen at minus 80°C, were thawed, equilibrated to room-temperature and then homogenized with a vortex for five seconds, and the 300 µl was mixed with 300 µl of NucliSENS® lysis buffer (Biomerieux, Marcy l’Etoile, France) and then extracted with the same procedure used for the nasopharyngeal samples. Saliva nucleic acids extracts were tested with the same RT-PCR procedure than for SARS-CoV-2 RT-PCR on nasopharyngeal samples.

#### RT-LAMP assay on saliva samples

The test EasyCov® (SkillCell-Alcen, Jarry, France) is a CE-marked extraction-free RT-LAMP test specifically developed for saliva samples. Detection of SARS-CoV-2 was carried out according manufacturer’s instructions (EasyCOV®, SkillCell) (appendix). In each screening center, saliva samples were tested immediately after collection (<5 minute) or stored immediately at 4°C and then tested within a maximal 90 minutes interval after collection. Briefly, 200 μl of saliva was mixed with the pretreatment buffer in tube 1 placed into the Easyvid® system for automated inactivation and lysis step (heating at 80 ° C for 10 minutes). Tube 1 is then taken out of Easyvid® and left to stand for 1 minute at room temperature. Three microliters of pretreated saliva sample are introduced into tube 2 containing RT-LAMP reaction mix. Tube 2 is incubated at 65 ° C for 30 minutes in dedicated positions of the Easyvid® system for viral RNA amplification. Once the 30 minutes have elapsed, tube 2 is taken out of the Easyvid® system and left to stand for 1 minute at room temperature. One microliter of revelation reagent is introduced in tube 2. The result is immediately read by visual observation. The color turns yellow for a sample positive for SARS-CoV2 RNA and remains orange for a sample negative for SARS-CoV2 RNA.

## References

1. Sharfstein JM, Becker SJ, Mello MM. Diagnostic Testing for the Novel Coronavirus. JAMA. 2020 Apr 21;323(15):1437–8.

2. Paltiel AD, Zheng A, Sax PE. Clinical and Economic Effects of Widespread Rapid Testing to Decrease SARS-CoV-2 Transmission. Ann Intern Med. 2021 Mar 9;

3. Bosetti P, Kiem CT, Yazdanpanah Y, Fontanet A, Lina B, Colizza V, et al. Impact of mass testing during an epidemic rebound of SARS-CoV-2: a modelling study using the example of France. Euro Surveill. 2021 Jan;26(1).

4. Du Z, Pandey A, Bai Y, Fitzpatrick MC, Chinazzi M, Pastore Y Piontti A, et al. Comparative cost-effectiveness of SARS-CoV-2 testing strategies in the USA: a modelling study. Lancet Public Health. 2021 Mar;6(3):e184–91.

5. Hanson KE, Caliendo AM, Arias CA, Englund JA, Lee MJ, Loeb M, et al. Infectious Diseases Society of America Guidelines on the Diagnosis of COVID-19. Clin Infect Dis. 2020 Jun 16;

6. Ricks S, Kendall EA, Dowdy DW, Sacks JA, Schumacher SG, Arinaminpathy N. Quantifying the potential value of antigen-detection rapid diagnostic tests for COVID-19: a modelling analysis. BMC Med. 2021 Mar 9;19(1):75.

7. Peeling RW, Olliaro PL, Boeras DI, Fongwen N. Scaling up COVID-19 rapid antigen tests: promises and challenges. Lancet Infect Dis. 2021 Feb 23;

8. Dinnes J, Deeks JJ, Berhane S, Taylor M, Adriano A, Davenport C, et al. Rapid, point-of-care antigen and molecular-based tests for diagnosis of SARS-CoV-2 infection. Cochrane Database Syst Rev. 2021 Mar 24;3:CD013705.

9. Takeuchi Y, Akashi Y, Kato D, Kuwahara M, Muramatsu S, Ueda A, et al. Diagnostic performance and characteristics of anterior nasal collection for the SARS-CoV-2 antigen test: a prospective study. Sci Rep. 2021 May 18;11(1):10519.

10. Lindner AK, Nikolai O, Rohardt C, Burock S, Hülso C, Bölke A, et al. Head-to-head comparison of SARS-CoV-2 antigen-detecting rapid test with professional-collected nasal versus nasopharyngeal swab. Eur Respir J. 2021 May;57(5).

11. To KK-W, Tsang OT-Y, Yip CC-Y, Chan K-H, Wu T-C, Chan JM-C, et al. Consistent Detection of 2019 Novel Coronavirus in Saliva. Clin Infect Dis. 2020 Jul 28;71(15):841–3.

12. Azzi L, Carcano G, Gianfagna F, Grossi P, Gasperina DD, Genoni A, et al. Saliva is a reliable tool to detect SARS-CoV-2. J Infect. 2020 Jul;81(1):e45–50.

13. Iwasaki S, Fujisawa S, Nakakubo S, Kamada K, Yamashita Y, Fukumoto T, et al. Comparison of SARS-CoV-2 detection in nasopharyngeal swab and saliva. J Infect. 2020 Aug;81(2):e145–7.

14. Tu Y-P, Jennings R, Hart B, Cangelosi GA, Wood RC, Wehber K, et al. Swabs Collected by Patients or Health Care Workers for SARS-CoV-2 Testing. N Engl J Med. 2020 Jul 30;383(5):494–6.

15. Lee RA, Herigon JC, Benedetti A, Pollock NR, Denkinger CM. Performance of Saliva, Oropharyngeal Swabs, and Nasal Swabs for SARS-CoV-2 Molecular Detection: A Systematic Review and Meta-analysis. J Clin Microbiol. 2021 Jan 27;

16. Butler-Laporte G, Lawandi A, Schiller I, Yao MC, Dendukuri N, McDonald EG, et al. Comparison of Saliva and Nasopharyngeal Swab Nucleic Acid Amplification Testing for Detection of SARS-CoV-2: A Systematic Review and Meta-analysis. JAMA Intern Med. 2021 Jan 15;

17. Bastos ML, Perlman-Arrow S, Menzies D, Campbell JR. The Sensitivity and Costs of Testing for SARS-CoV-2 Infection With Saliva Versus Nasopharyngeal SwabsC: A Systematic Review and Meta-analysis. Ann Intern Med. 2021 Jan 12;

18. Fernández-González M, Agulló V, Rica A de la, Infante A, Carvajal M, García JA, et al. Performance of saliva specimens for the molecular detection of SARS-CoV-2 in the community setting: does sample collection method matter? Journal of Clinical Microbiology [Internet]. 2021 Jan 8 [cited 2021 Mar 16]; Available from: https://jcm.asm.org/content/early/2021/01/08/JCM.03033-20

19. Kernéis S, Elie C, Fourgeaud J, Choupeaux L, Delarue SM, Alby M-L, et al. Accuracy of antigen and nucleic acid amplification testing on saliva and naopharyngeal samples for detection of SARS-CoV-2 in ambulatory care. medRxiv. 2021 Apr 11;2021.04.08.21255144.

20. Wolters F, van de Bovenkamp J, van den Bosch B, van den Brink S, Broeders M, Chung NH, et al. Multi-center evaluation of cepheid xpert® xpress SARS-CoV-2 point-of-care test during the SARS-CoV-2 pandemic. J Clin Virol. 2020 Jul;128:104426.

21. Subsoontorn P, Lohitnavy M, Kongkaew C. The diagnostic accuracy of isothermal nucleic acid point-of-care tests for human coronaviruses: A systematic review and meta-analysis. Sci Rep. 2020 Dec 18;10(1):22349.

22. Chen JH-K, Yip CC-Y, Poon RW-S, Chan K-H, Cheng VC-C, Hung IF-N, et al. Evaluating the use of posterior oropharyngeal saliva in a point-of-care assay for the detection of SARS-CoV-2. Emerg Microbes Infect. 2020 Dec;9(1):1356–9.

23. Nagura-Ikeda M, Imai K, Tabata S, Miyoshi K, Murahara N, Mizuno T, et al. Clinical Evaluation of Self-Collected Saliva by Quantitative Reverse Transcription-PCR (RT-qPCR), Direct RT-qPCR, Reverse Transcription-Loop-Mediated Isothermal Amplification, and a Rapid Antigen Test To Diagnose COVID-19. J Clin Microbiol. 2020 Aug 24;58(9).

24. Yamazaki W, Matsumura Y, Thongchankaew-Seo U, Yamazaki Y, Nagao M. Development of a point-of-care test to detect SARS-CoV-2 from saliva which combines a simple RNA extraction method with colorimetric reverse transcription loop-mediated isothermal amplification detection. J Clin Virol. 2021 Mar;136:104760.

25. Taki K, Yokota I, Fukumoto T, Iwasaki S, Fujisawa S, Takahashi M, et al. SARS-CoV-2 detection by fluorescence loop-mediated isothermal amplification with and without RNA extraction. J Infect Chemother. 2021 Feb;27(2):410–2.

26. L’Helgouach N, Champigneux P, Schneider FS, Molina L, Espeut J, Alali M, et al. EasyCOVC: LAMP based rapid detection of SARS-CoV-2 in saliva. medRxiv. 2020 May 30;2020.05.30.20117291.

27. Marot S, Calvez V, Louet M, Marcelin A-G, Burrel S. Interpretation of SARS-CoV-2 replication according to RT-PCR crossing threshold value. Clin Microbiol Infect. 2021 Jan 29;

28. Bossuyt PM, Cohen JF, Gatsonis CA, Korevaar DA, STARD group. STARD 2015: updated reporting guidelines for all diagnostic accuracy studies. Ann Transl Med. 2016 Feb;4(4):85.

29. Santos Schneider F, Molina L, Picot MC, L’Helgoualch N, Espeut J, Champigneux P, et al. Comparative Evaluation of Rapid Salivary RT-LAMP Assay for Screening of SARS-CoV-2 Infection [Internet]. Rochester, NY: Social Science Research Network; 2021 Feb [cited 2021 Jun 2]. Report No.: ID 3774184. Available from: https://papers.ssrn.com/abstract=3774184

30. Yokota I, Shane PY, Okada K, Unoki Y, Yang Y, Iwasaki S, et al. A novel strategy for SARS-CoV-2 mass screening with quantitative antigen testing of saliva: a diagnostic accuracy study. The Lancet Microbe [Internet]. 2021 May 19 [cited 2021 Jun 2];0(0). Available from: https://www.thelancet.com/journals/lanmic/article/PIIS2666-5247(21)00092-6/abstract

